# Perceptions and Practices Among Students, Parents, and Teachers of the Human Papilloma Virus( HPV) vaccine : A Pilot Feasibility Study from Rural Kerala

**DOI:** 10.64898/2025.12.25.25343008

**Authors:** Anjum John, Roshna Rasheed, Mercy John Idikula

## Abstract

**Background:** India bears the world’s highest cervical cancer burden, yet awareness and uptake of the effective HPV vaccine remain low. Although Kerala has initiated pilot HPV vaccination programmes, stakeholder knowledge, attitudes, and practices (KAP) are not well documented. This pilot study aimed to assess the feasibility of a KAP study and to identify gaps in the questionnaire, for future analytical modelling.

**Methods:** A cross-sectional pilot study was conducted in a school in Pulinkunnu, Kerala, among students aged 11–15 years, their parents, and teachers. A structured, validated questionnaire, developed in English, translated into Malayalam, was administered during school hours. Responses were entered into secure electronic spreadsheets. Descriptive statistics summarised KAP outcomes. As key demographic and behavioural predictors—such as gender, socioeconomic status, prior health education, and exposure to HPV information—were not captured, multivariable analysis was not undertaken.

**Results:** The pilot included 75 students and 20 teachers, predominantly female. HPV-related knowledge was low across groups, with widespread misconceptions and limited awareness of vaccine availability and cervical cancer screening. Attitudes toward vaccination were mixed, marked by safety concerns and modest willingness to receive or recommend the vaccine. Practices reflected low prior awareness and uptake, with few respondents having heard of or received the HPV vaccine.

**Conclusions:** This pilot revealed major gaps in HPV-related knowledge, attitudes, and practices, among stakeholders and highlighted the need for an improved questionnaire in item clarity, response patterns, and missing variables and a larger study to identify predictors and inform targeted school-based HPV education efforts.

**Ethics and Dissemination:** Ethical approval for this pilot study was obtained from the Institutional Ethics Committee of Pushpagiri Institute of Medical Sciences and Research Centre. Written informed consent was obtained from participating teachers, and parental consent was sought for students; however, parents were unavailable on the day of data collection, resulting in their non-participation. Verbal assent was obtained from all student participants prior to survey administration.

**Patient and Public Involvement:** Students, teachers, and parents were not involved in the development of the research protocol or the pilot questionnaire. As this was a preliminary feasibility study, no direct involvement in interpreting or disseminating the results is planned at this stage. Insights from the pilot will instead be used internally by the research team to refine the study tools and procedures for the full-scale investigation.

## Introduction

Human Papillomavirus (HPV) is the most common sexually transmitted infection globally, with around 40 strains associated with infections of the anogenital region. Although most of these infections are self-limiting, certain high-risk strains are linked to cancers such as cervical, anal, oropharyngeal, and penile cancer. (1)Despite the availability of effective vaccines, HPV-related diseases remain a major public health concern, particularly in low- and middle-income countries where screening and vaccination coverage are limited. Understanding public perspectives and practices (KAP) regarding HPV and its vaccine is therefore critical to shaping effective prevention strategies.(2-5)

### Public health burden of HPV

Globally, cervical cancer is the fourth most common cancer among women. In 2023, it was estimated that there were 604,000 new cervical cancer cases, causing around 342,000 deaths worldwide. Projections suggest an annual increase of nearly 100,000 new cases between 2018 and 2030. Prevalence of HPV is influenced by social and economic determinants like gender inequality and poverty, with illiteracy and early sexual initiation contributing to over 85% of cervical cancer cases, with an additional risk in people living with Human Immunodeficiency virus ( HIV) infections.(6, 7) The disease threatens the well-being of children when mothers die prematurely due to cervical cancer.(8)

### Disparities in healthcare

Cervical cancer is a preventable and curable disease. Despite the availability of the HPV vaccine for the prevention of cervical cancer, and effective treatment measures-much of the world has limited access to these services. Deaths due to cervical cancer are three times higher in low- or lower-middle-income countries, demonstrating the widespread disparities in the availability, accessibility, and adaptability of health care services in regions with the highest disease burden.(9)

Integrating HPV vaccination, cervical cancer screening, and management of precancerous lesions/cervical cancer into health systems remain essential. Until then, cervical cancer will continue to be one of the leading causes of cancer death in women. Involving people in their own healthcare helps reduce the incidence, mortality, and burden of HPV-related conditions. (10)The demonstrated success of HPV vaccination has prompted a global commitment to eliminate cervical cancer as a public health problem. (11)

### Global strategy

Public health microbiology is at the threshold of a rare but doable feat- the elimination of a non-communicable disease-cervical cancer.The World Health Organization’s 2020 Global Strategy to eliminate cervical cancer set the 90-70-90 targets by 2030: vaccinate 90% of girls under 15 years, screen 70% of women at 35 and 45 years of age, and treat 90% of women identified with cervical disease. (7, 11)

### HPV in India

India has the highest global burden of cervical cancer. Annually, about 132,000 new cases and 80,000 deaths occur, with a high prevalence of HPV type 16.(5, 6)The vastness of the country and the diversity of its population are responsible for the lack of proper documentation of the epidemiology and patterns of HPV virus strains. (10) Thus, as in other lower income countries, the implementation of prevention and control programs has been difficult. This highlights the importance of HPV vaccination in India.

### Vaccine coverage in India and Kerala

India licensed two HPV vaccines in 2008. The state of Sikkim successfully implemented the first school-based immunization program in 2018, achieving >95% coverage with strong government and community support. (12)In 2022, the National Technical Advisory Committee on Immunization recommended introducing the indigenously developed quadrivalent vaccine into the Universal Immunization Program (UIP). The vaccine rollout was planned for 2024, with Kerala’s Wayanad and Alappuzha districts included in the pilot phase.(13-16)

In Kerala, voluntary uptake remains poorly documented, and some studies report declining cervical cancer rates (7–9/100,000 women annually), which has sometimes been used to argue for screening rather than vaccination. (17) A 2018 study found only 8% coverage among medical students. (18) More recently, the Happy Noolpuzha Initiative in Wayanad demonstrated local-level success by vaccinating tribal girls through a Panchayat-led campaign.(19)

### Review of literature

Prior studies in India show low levels of HPV awareness and vaccination. Backer et al. (2024) found that although 85.6% of college students in Kerala knew HPV causes cervical cancer, only 48.6% had heard of the vaccine, and uptake was low (11.6% in medical students, 4.9% in non-medical).(20) Basu et al. similarly reported limited uptake among adolescents in North India, citing lack of awareness, cost, and cultural hesitancy. (21)School-based vaccination and awareness campaigns have proven effective in increasing acceptance.(22-24) However, teachers and parents often lack adequate knowledge, with safety concerns and stigma acting as major barriers. (25-27) Our full-scale study aimed to fill this gap by assessing stakeholder knowledge and perceptions in the Kerala context.

### Rationale for the present study

Limited evidence exists on the knowledge, attitudes, and practices of school-going adolescents, parents, and teachers—key stakeholders in vaccine acceptance. This gap limits the development of effective, school-based prevention strategies, despite ongoing HPV vaccination initiatives. Additionally, no validated tools were available to assess HPV-related knowledge and perceptions in this setting, necessitating the development and pilot testing of a new questionnaire. This pilot study was therefore undertaken to evaluate the feasibility of the tool, study item clarity and completeness of variables, and ensure that the instrument could adequately capture stakeholder perspectives before implementing a full-scale study. By identifying gaps in awareness, acceptance, and questionnaire performance, this pilot aimed to generate preliminary insights to guide a larger, more comprehensive investigation. The protocol for this study was published in the Indian Journal of Applied Research in October 2025.( doi: 10.36106/ijar and ISSN No : 2249-555X available from (https://www.worldwidejournals.com/indian-journal-of-applied-research-(IJAR)/fileview/knowledge-attitude-and-practice-regarding-human-papillomavirus-hpv-vaccine-among-school-students-parents-and-teachers-in-central-kerala-a-crosssectional-study-protocol_October_2025_5111739822_7103697.pdf)

### Objectives

The study was conducted to assess the feasibility of measuring knowledge, attitudes, and practices related to HPV and HPV vaccination among school students aged 11–15 years, their teachers, and their parents, in preparation for a larger study.

## Methods

### Study design and setting

A pilot cross-sectional study was conducted in 2025 at a higher secondary school in Pulinkunnu, Alappuzha, Kerala, targeting students aged 11–15 years, their teachers, and their parents. This study was explicitly designed as a pilot to refine instruments, assess feasibility, and inform changes to the questionnaire and resource scale up, consistent with recommended guidelines for pilot and feasibility studies. (28, 29)

### Study participants

All students aged 11–15 years enrolled in the selected school and present on the day of data collection were eligible to participate, subject to informed assent and parental consent. Teachers of the selected classes and parents of enrolled students were also invited to participate.

### Sample size and sampling

As this was a pilot study, universal sampling was employed. All students aged 11–15 years enrolled in the selected school and present on the day of data collection were invited to participate. All teachers responsible for these classes were also included. Parents of enrolled students were invited to complete the questionnaire.

#### Tools

A brief structured questionnaire was developed with close-ended questions to assess the perceptions and practices related to HPV infection and vaccination. Given the exploratory objectives of the pilot, sociodemographic information was not collected. A single, common questionnaire was used for both parents and teachers to ensure uniformity and reduce respondent burden. Questionnaire items were informed by a review of existing KAP studies on HPV vaccination and were framed using simple, non-judgmental language to enhance comprehension and participation. Content validity was assessed through review by subject experts from Community Medicine and Biostatistics, who evaluated each item for relevance, clarity, and appropriateness for the target population. Face validity and feasibility were further examined through pilot testing among a small group of postgraduate students and subject experts, focusing on clarity, length, and ease of answering questions. Feedback from this process led to minor refinements, primarily aimed at shortening the questionnaire and improving wording.

Internal consistency reliability of the final tool was assessed using Cronbach’s alpha, which demonstrated acceptable reliability for the overall questionnaire (Cronbach’s α ≥0.7), supporting its suitability for use in the pilot study.

### Data Collection

A structured questionnaire was developed in English, translated into Malayalam, and validated for content and face validity. Trained investigators administered the survey to students and teachers during school hours using paper-based forms. Parents were provided questionnaires through their wards. All completed forms were anonymized, coded, and entered into secure, password-protected electronic spreadsheets.

### Recruitment Period

Data was collected between the 18^th^ and 19^th^ of June, 2025. Parental consent was taken on the first day, with the questionnaires and information sheet about the research sent home with the students for parental review. Student assent was taken on the second day before the questionnaires were filled in by them. Teacher consent was taken during the day of the study.

### Outcome Measures

The pilot study assessed the feasibility and clarity of items measuring knowledge of HPV and cervical cancer, awareness of the HPV vaccine, attitudes toward vaccination, willingness to receive or recommend the vaccine, perceived barriers, vaccination-related practices, and sources of information. These outcomes were used primarily to evaluate the performance of the questionnaire and identify areas requiring refinement for the follow up full-scale study.

### Data analysis

Data were cleaned and analysed using descriptive statistics to summarize sociodemographic characteristics and KAP outcomes. With no key demographic or behavioural variables available for multivariable modelling, no inferential or regression analyses were undertaken. The focus of analysis was to evaluate response patterns, identify missing or non-discriminating items, and assess the overall feasibility and suitability of the questionnaire for use in a larger study.

## Results

### Demographic Profile

The study was conducted at an Indian Certificate of School Education(ICSE) school at Pulinkunnu and included 75 students and 20 teachers. No parents participated, as no student returned the parent questionnaire, representing a deviation from the planned study design. Among the students, 16 (21.3%) were boys and 59 (78.7%) were girls. Among teachers, 2 (10%) were male and 18 (90%) were female. The majority of students were in the 10–12-year age group (45.3%), followed by 13–15 years (36%) and 16–17 years (18.7%). Most teachers were between 40–49 years (40%), with equal proportions in the 30–39 years and 50–59 years categories (30% each).

### 1. Knowledge Interpretation ( Table 2)

#### Low levels of knowledge of HPV and cervical cancer linkage

Only about half of students (50.7%) and teachers (45%) correctly identified that HPV causes cervical cancer. This indicates that foundational knowledge of HPV was inadequate in this school community.

**Table 1:** Age classification of the students and teachers.

| Age Group | Frequency | Percentage |
| --- | --- | --- |
| 10-12 years | 34 | 45.3% |
| 13-15 years | 27 | 36% |
| 16-17 years | 14 | 18.7% |
|  | **75** |  |
| Teachers |  |  |
| 30-39 years | 6 | 30% |
| 40-49 years | 8 | 40% |
| 50-59 years | 6 | 30% |
|  | **20** |  |

**Table 2:**
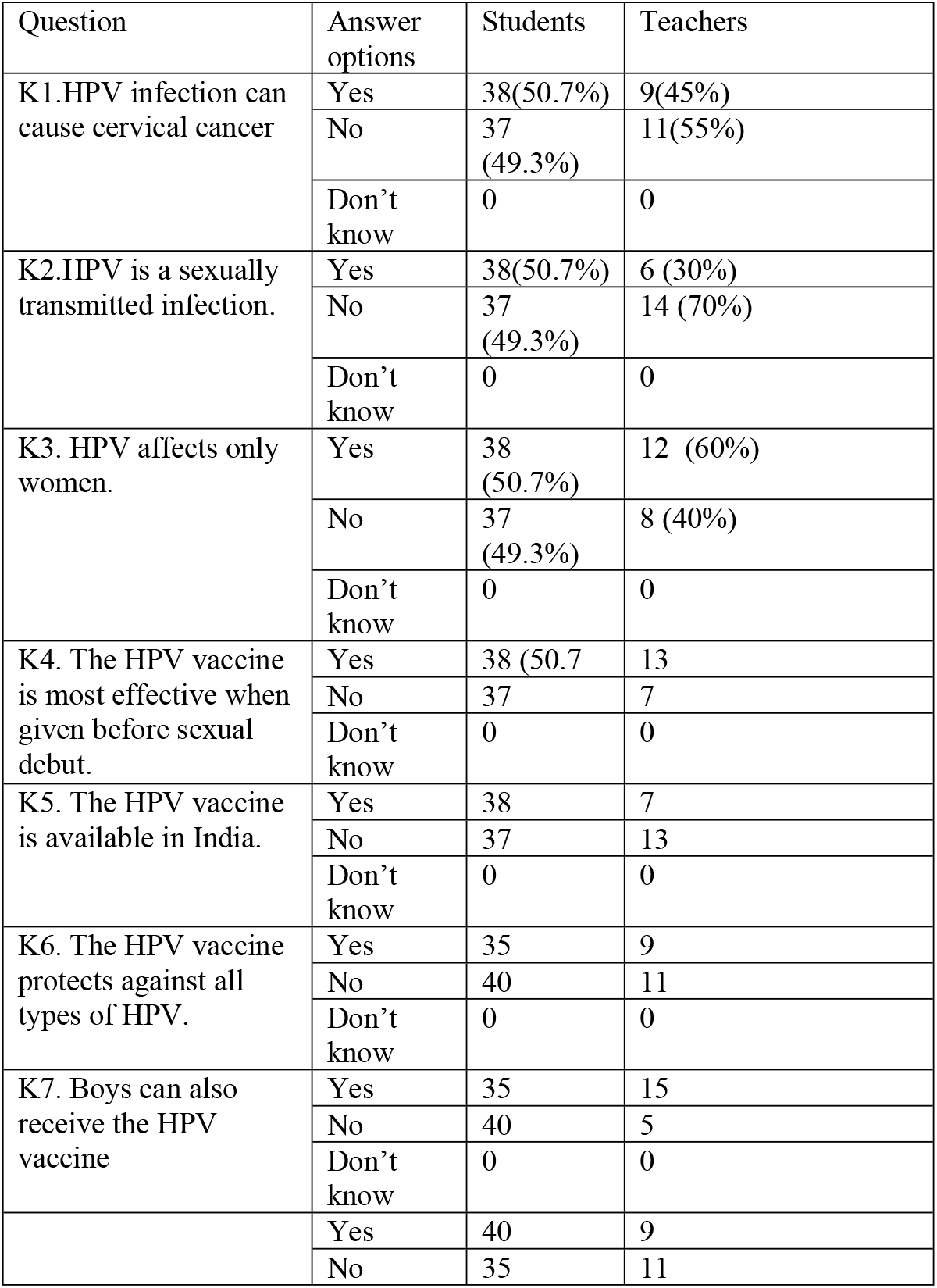
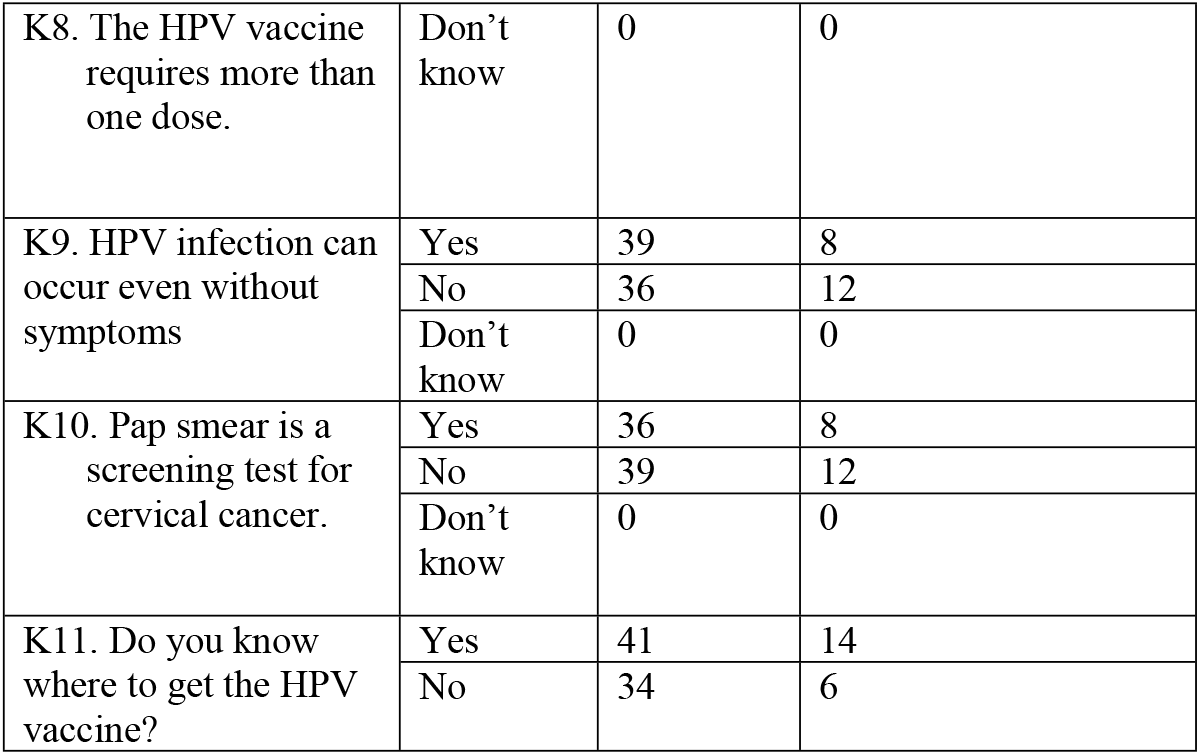
Knowledge about HPV vaccines.

#### Poor understanding of HPV transmission

Only 30% of teachers and half the students correctly identified HPV as a sexually transmitted infection. This was important because misunderstanding transmission pathways could weaken risk perception and minimise urgency for vaccination.

#### Significant misconceptions

Half the students and 60% of teachers incorrectly believed HPV affects only women. Almost half did not know the vaccine was available in India. Many believed HPV infection always presented with symptoms. These misconceptions implied that health education on HPV in schools had not been effectively integrated, and teachers themselves required orientation before offering student guidance.

#### Limited knowledge of screening methods

Only 36 students and 8 teachers knew Pap smear was used for cervical cancer screening. This suggested major gaps in understanding of the prevention continuum (screening + vaccination) for HPV cervical cancer.

#### Better awareness about vaccination eligibility for boys

Students and teachers showed comparatively better awareness that boys can receive HPV vaccines. This was encouraging and aligning with current recommendations.

Overall, knowledge levels were low-to-moderate, with several misunderstandings that could impede vaccine uptake.

### 2. Attitude Interpretation( Table 3)

#### “Neutral” and “hesitant” attitudes dominate

Most respondents did not strongly support HPV vaccination; neutral responses were common across nearly all attitude items. This indicated uncertainty rather than rejection.

**Table 3:** Attitude about HPV Vaccines.

| Attitude Questions | Answer Options | Students | Teachers |
| --- | --- | --- | --- |
| A1. HPV vaccination should be included in the national immunization schedule. | Strongly Agree | 15 | 2 |
|  | Agree | 15 | 6 |
|  | Neutral | 19 | 4 |
|  | Disagree | 15 | 4 |
|  | Strongly Disagree | 11 | 4 |
| A2. I believe HPV vaccination is important for cancer prevention. | Strongly Agree | 11 | 2 |
|  | Agree | 12 | 4 |
|  | Neutral | 16 | 3 |
|  | Disagree | 15 | 5 |
|  | Strongly Disagree | 21 | 6 |
| A3. I would recommend the HPV vaccine to others. | Strongly Agree | 15 | 8 |
|  | Agree | 18 | 4 |
|  | Neutral | 10 | 3 |
|  | Disagree | 17 | 2 |
|  | Strongly Disagree | 14 | 3 |
| A4. I worry about the side effects of the HPV vaccine. | Strongly Agree | 19 | 3 |
|  | Agree | 14 | 8 |
|  | Neutral | 17 | 3 |
|  | Disagree | 10 | 2 |
|  | Strongly Disagree | 15 | 4 |
| A5. HPV vaccination promotes early sexual activity | Strongly Agree | 13 | 4 |
|  | Agree | 11 | 2 |
|  | Neutral | 20 | 5 |
|  | Disagree | 11 | 2 |
|  | Strongly Disagree | 20 | 7 |
| A6. I believe awareness campaigns are needed for HPV vaccination. | Strongly Agree | 14 | 1 |
|  | Agree | 13 | 3 |
|  | Neutral | 17 | 8 |
|  | Disagree | 13 | 5 |
|  | Strongly Disagree | 18 | 3 |
| A7. I trust the safety of the HPV vaccine. | Strongly Agree | 14 | 1 |
|  | Agree | 13 | 3 |
|  | Neutral | 17 | 8 |
|  | Disagree | 13 | 5 |
|  | Strongly Disagree | 18 | 3 |
| A8. If not vaccinated, are you willing to take it? | Yes | 33 | 8 |
|  | No | 42 | 12 |

**Table 4:**
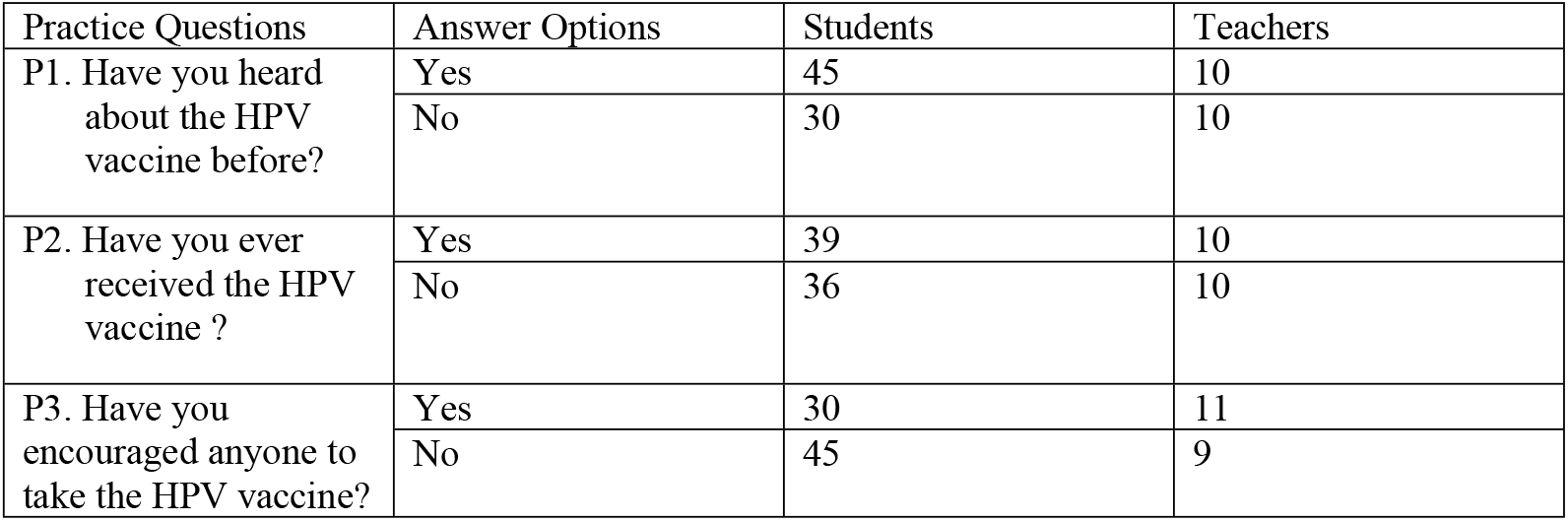
Practice about HPV Vaccines.

| Practice Questions | Answer Options | Students | Teachers |
| --- | --- | --- | --- |
| P1. Have you heard about the HPV vaccine before? | Yes | 45 | 10 |
|  | No | 30 | 10 |
| P2. Have you ever received the HPV vaccine ? | Yes | 39 | 10 |
|  | No | 36 | 10 |
| P3. Have you encouraged anyone to take the HPV vaccine? | Yes | 30 | 11 |
|  | No | 45 | 9 |

#### Concerns about side effects were high

Around one-third strongly agreed/agreed that they worried about vaccine side effects ( Question A4).

#### Belief that HPV vaccination may promote early sexual activity

Approximately 32% of students and teachers agreed or strongly agreed with this misconception. This reflected socio-cultural norms and moral concerns, which have historically influenced vaccine hesitancy.

#### Mixed trust in vaccine safety

Many respondents remained neutral or disagreed that HPV vaccines were safe. This suggested the need for targeted communication addressing safety, effectiveness, and global evidence of long-term benefit.

#### Willingness to be vaccinated

Despite low knowledge levels, 44% of students and 40% of teachers expressed willingness to take the vaccine if not already vaccinated. This indicated that improving awareness could translate into greater uptake.

From the responses to the “Attitude” questions, there seems to be a “hesitant to vaccine-uptake” population, where attitude shifts through education are required.

### 3. Practice Interpretation

#### Moderate awareness but low actual vaccination

While 45 students and 10 teachers had heard of HPV vaccines, only 39 students and 10 teachers had received it.

This mismatch suggested barriers to access vaccines, low parental support, safety concerns about the HPV vaccine, and limited communication from schools or healthcare providers due to ‘vaccine shyness’, or sociocultural factors that prevent them from talking about the vaccine.

#### Low rates of recommending vaccination to others

Only 30 students and 11 teachers reported encouraging someone else to take the vaccine.

This aligned with their low confidence and incomplete knowledge, possibly reflecting the unsurety of individuals about the HPV vaccine safety and necessity.

#### Absence of parental participation

No parents participated in the survey . This highlighted difficulty involving parents—who are the key decision-makers for adolescent vaccination.

Overall, HPV vaccine practices are poor, reflecting the knowledge deficits and hesitant attitudes and low practice parameters.

#### Demographic Interpretation

Majority of students were girls, which might have influenced responses as cervical cancer is often perceived as a “female-only” issue. Teachers were predominantly female and in mid-career age groups; traditionally such groups were expected to support adolescent health programs, yet knowledge levels among this cohort were low. This suggested the need for structured teacher training before implementing school-based HPV initiatives.

Insights from the pilot study to guide a scale-up study

Though levels of knowledge, attitudes, and self-reported practices about the HPV vaccine could be described, the authors realised the study was unable to identify meaningful predictors of good knowledge, positive attitudes, or favourable practices related to HPV vaccination, particularly those operating at the parental, household, school, and socioeconomic levels as these variables were either absent or inadequately captured.

From a feasibility perspective, the pilot demonstrated that though KAP studies are good to capture the reasons behind health decisions at a point in time, not capturing contextual and decision-making variables limits interpretability. Parent and teacher practices around HPV vaccination are likely influenced by complex social, cultural, and informational factors that are not easily elicited through closed-ended questions. Interview-based or mixed-methods approaches may be necessary to understand how adults interpret vaccine information, perceive risk, and make decisions for adolescents. These insights underscored the need to reframe the questionnaire and to incorporate qualitative methods in the main study.

## Discussion

This school-based study provides critical insights into the knowledge, attitudes, and practices related to HPV vaccination among students and teachers. The findings reveal substantial gaps in awareness, widespread misconceptions, and low vaccination uptake despite moderate exposure to HPV-related information. When examined alongside the national and global evidence base, the results highlight systemic information gaps, sociocultural influences, and structural barriers that continue to limit HPV vaccine acceptability and uptake in India.

### Knowledge Levels Compared with Other Studies

In this study, only about half of students and teachers recognised that HPV causes cervical cancer. These findings align with several Indian studies demonstrating poor foundational knowledge about HPV. Backer et al. (2024) reported that although 85.6% of Kerala college students knew HPV was linked to cervical cancer, only 48.6% had heard of the vaccine.(20) Similar patterns were seen in North India, where Basu et al. found that adolescents were largely unaware of HPV transmission and its association with cervical cancer. (21) Internationally, limited awareness is also well-documented in Low Middle-Income Countries( LMIC), particularly where sexual health education is restricted. (30, 31)

A major misconception observed was the belief that HPV affects only women. This misconception has been highlighted globally. Studies from Europe and the United States show that boys often underestimate their susceptibility to HPV-related diseases, including penile, oropharyngeal, and anal cancers. (32, 33) This gendered perception reduces motivation for male vaccination, despite WHO and CDC recommendations for universal vaccination of both sexes. (34-36)

Similarly, inadequate awareness that the HPV vaccine is available in India reflects a lack of systematic public communication. Many studies have found that even medical students were uncertain about vaccination availability and access points. (37, 38) In contrast, Sikkim’s school-based programme achieved high awareness due to intensive pre-implementation education. (12) The contrast indicates that Kerala has not yet conducted similar structured outreach.

Overall, the knowledge gap in our study is consistent with national trends and highlights the absence of integrated HPV education in schools, even in high-literacy states like Kerala.

### Misconceptions and Underlying Influences

Fear of HPV vaccine side effects was prominent in our study, echoing findings from several Indian and global studies. Seo et al. reported through their scoping review that concerns about safety and adverse events are the leading reason for HPV vaccine refusal among parents.(39) Asmelash et al. similarly identified safety concerns as a major determinant of vaccine hesitancy across West and East African countries. (40)Such concerns are often fuelled by misinformation, lack of transparent communication, and limited exposure to scientific evidence.

Another recurring misconception was the belief that HPV vaccination may promote early sexual activity. This has been repeatedly shown to be untrue in scientific literature, yet remains a widespread cultural fear. Studies in Turkeye, Africa, and South Korea report that parents worry vaccination may be interpreted as permission for sexual activity.(41-43) Studies from the US, however, show no association between HPV vaccination and sexual behaviour . (44)The persistence of this misconception highlights cultural discomfort with sexuality education, particularly in conservative settings.

Misconceptions about the number of doses required and uncertainty about asymptomatic HPV infections further indicate the absence of structured, accurate health communication in schools. Similar misunderstandings have been reported in East Asian and African countries, where lack of clarity about vaccine schedules hinders adherence. (42, 45-47)

### Attitudes Toward HPV Vaccination in Context

The predominance of “neutral” or “uncertain” attitudes in our study indicates “ambivalent” hesitancy rather than outright refusal. This is widely observed in vaccine acceptance research. Lelliott et al. describe ambivalence as a form of “information hesitancy,” where individuals are unsure due to incomplete knowledge rather than opposition.(48) Backer et al. and Muthukrishnan et al. also report that unclear, inconsistent, or conflicting information contributes to passive resistance to vaccination.(20, 49)

Safety concerns were common in our sample, consistent with a recent study by Waheed et al., and an early study by Holman, which identified perceived risk of adverse events as the most influential barrier to HPV vaccination globally. (50, 51) Conversely, countries with long-standing HPV vaccination programmes—such as Australia and the United Kingdom—report significantly lower safety-related hesitancy because of strong public trust and robust adverse event monitoring. (52, 53)

Importantly, willingness to be vaccinated was relatively high once individuals were informed, indicating that hesitancy is modifiable. Aslemash et al. demonstrated that even brief educational interventions significantly increase willingness to vaccinate. (40)Similar improvements have been documented in US and European settings. (54) Thus, the results of our study suggest that structured school-based awareness programmes could rapidly improve acceptance in Pulinkunnu.

### Practices and the Knowledge–Behaviour Gap

Despite moderate awareness, actual vaccination uptake remained low among both students and teachers. This gap between intention and practice is well documented in India. Basu et al. found that even when adolescents expressed interest, parental concerns, cost, and lack of access remained barriers. (21) Das et al. reported low uptake even among medical students, demonstrating that knowledge in some settings does not directly translate to action.(18)

The absence of parental participation in our study further highlights a major barrier. Numerous studies have shown that parental consent and beliefs are the strongest predictors of adolescent HPV vaccination. In South India, for example, parental attitudes accounted for more than 50% of vaccination decisions. (27, 55)The complete absence of parental responses here underscores the urgent need to involve families in HPV awareness initiatives.

When compared with successful models, the gap becomes clearer. Sikkim’s programme and the Happy Noolpuzha campaign in Wayanad achieved high coverage through community mobilisation, strong local leadership, and parent-centred communication. (12, 19) Our findings suggest that without such community engagement, school-based programmes alone may be insufficient.

### Proposed Changes to existing questionnaires in preparation for future research

#### a. Add variables that can act as predictors

Sociodemographic predictors like parental level of education, occupation and socioeconomic status, family structure and decision-maker for child vaccination in families, and prior exposure to health campaigns or school health programs are all important predictors which must be studied. Specific health system and information exposure questions need be added like source of HPV vaccine information, level of trust in healthcare providers and government programs, previous experience with childhood or adolescent vaccinations, or awareness of government HPV vaccination policies. Cultural and belief-based factors are another dimension to be explored like perceived stigma around sexually transmitted infections, comfort discussing sexual health topics, perceived moral or cultural concerns related to HPV vaccination, gender norms influencing adolescent health decisions.

#### b. Modify data-collection mode

The pilot indicated that self-administered online questionnaires may not be optimal, particularly for parents, as literacy and digital familiarity may affect response quality. The following methodological changes are being proposed : interviewer-administered, short, semi-structured questionnaires for parents and teachers where possible to explore reasoning behind practices and capture unanticipated concerns or motivations. These would enhance data completeness, depth, and validity.

##### Public Health Implications

The findings of this study highlight the requirement for three strategic actions. School based health education with structured sessions for teachers, parents, and students addressing safety, immunisation schedules, benefits, costs, adverse events, and availability of the HPV are required. Parental involvement with regular meetings, and open house sessions with educational brochures for clarifying doubts are recommended. The places where HPV vaccines are available, the methods to procure the vaccine etc should be provided to ensure accessibility and clarity for referral. These strategies align with WHO’s elimination framework and the national plan for HPV vaccination under the Universal Immunisation Program( UIP).

## Strengths of the pilot study

A key strength of this pilot study lies in its feasibility-driven design. Conducted as a preliminary investigation, the study was essential for refining the student, teacher, and parent questionnaires, particularly to identify and restructure items capturing key predictor variables for HPV vaccine knowledge, attitudes, and acceptance. The use of a validated questionnaire, combined with insights gained from pilot testing, enhanced content clarity, contextual relevance, and construct coverage for the main study.

The pilot study conducted among 75 students and 20 teachers in a socio-demographically comparable setting—the neighbouring district of Alappuzha( district neighbouring to Pathanamthitta district where the full study is planned)—allowed assessment of recruitment processes, participation rates, and data-collection logistics without contaminating the main study population. The pilot also provided realistic estimates of time, personnel, and material resources required for large-scale implementation, thereby strengthening the methodological preparedness of the primary study.

Limitations include the absence of parent responses during the pilot phase, the single-school setting, and reliance on self-reported practices, which may be subject to reporting bias. Online or self-filled formats (e.g., Google Forms) were associated with incomplete responses, superficial answers, and difficulty probing reasoning behind decisions.

However, consistent with the purpose of pilot and feasibility studies, the findings are not intended to be generalisable but to inform study design and implementation. The study nevertheless provides valuable baseline insights from a rural context relevant to HPV vaccine introduction.

## Conclusion

The study demonstrates major knowledge gaps, mixed attitudes, and low HPV vaccination uptake among students and teachers, though willingness improves when informed. Strengthening school-based awareness, involving parents, and improving access to vaccination services will be critical for successful programme implementation. With national HPV rollout underway, these findings can guide local action for cervical cancer prevention.

## Data Availability

The data cannot be shared publicly because of privacy reasons but they are available with the authors and will be provided anonymously, upon reasonable request.

## Key findings

| Domain | Observations | Implication |
| --- | --- | --- |
| Knowledge | ~50% knew HPV causes cervical cancer | Large gaps despite high literacy |
| Misconceptions | HPV affects only women; vaccine not available | Targeted correction needed |
| Attitudes | Neutral or uncertain responses dominant | Hesitancy rather than rejection |
| Safety Concerns | Common among both groups | Need for counselling |
| Uptake | Low vaccination despite willingness | Access and family involvement critical |

